# Real-Time Genomic Epidemiology Approaches to a Measles Outbreak Response in Utah

**DOI:** 10.64898/2026.08.25.26361356

**Authors:** Mary Jewell, Abbey Marye, Clarissa Nielsen, Leisha D. Nolen, Amelia Salmanson, Kelly Oakeson

**Author notes:** These authors contributed equally to this work (Mary Jewell, Abbey Marye). Author order was chosen randomly.

## Abstract

As vaccination rates have declined, large measles outbreaks have taken hold in vulnerable populations. In 2025, the United States saw the highest number of measles cases since 1991. After Utah’s first case in June 2025, the Utah Public Health Laboratory (UPHL) began performing whole genome sequencing (WGS) on clinical measles samples. As Utah’s outbreak grew, public health professionals used WGS data to identify clusters of disease, and combined genomic and epidemiologic data to identify factors that may fuel the spread of disease throughout the state. This study used sequenced samples from 66% of reported measles cases in Utah. Time-scaled and maximum likelihood phylogenetic trees were generated. Genomic-based transmission clusters were created using a transmission threshold of eight and an R_0_ of five. These clusters were used in a logistic regression to determine the association between genomic cluster and school affiliation. Epidemiological clusters were assessed using annotated phylogenetic trees. We found multiple introductions of measles into Utah, with one accounting for the majority of cases. As measles spread, two phylogenetic clades emerged with differing geographical case compositions. We identified a significant association between shared school and genomic clustering, and we reconstructed transmission chains within schools and emerging from school sporting events. This study demonstrates the utility of WGS in real-time outbreak investigations. Sequencing data allowed for gaps in epidemiological data to be filled, revealing undetected transmission and clarifying whether cases belong to a known outbreak. We also highlight that schools and high-contact sporting events played a significant role in fueling transmission across the state.

**Importance:** As vaccination rates decline, measles outbreaks will continue to appear across the United States. WGS has become easier to integrate into public health laboratories, as has genomic data into epidemiological workflows. Due to the nature of measles outbreaks, WGS is a valuable tool to fill in the gaps of epidemiological investigations and assess potential areas or events at risk of greater transmission. In this study, we show that utilizing WGS in real time was crucial in our understanding of how transmission occurred following large gatherings. By demonstrating the utility of integrating WGS into a real-time epidemiological workflow on a large scale, this study provides a launching point for further application of genomic data to investigate and understand emerging outbreaks.

## Background

Measles is a highly contagious respiratory disease that was once considered a common childhood illness, infecting between 3–4 million people and causing 400–500 deaths in the United States annually before the adoption of the vaccine (1). Since the vaccine was introduced, the annual incidence of measles dropped substantially, and it was declared eliminated in the United States in 2000 (1,2). However, because the measles virus is highly infectious and can spread rapidly through airborne and droplet transmission, it requires vaccination coverage between 96.1-97.8% to reach herd immunity (3). Achieving this level of vaccination coverage can be difficult. Due to its high infectivity, measles becomes difficult to track with traditional epidemiological investigations, particularly in larger outbreaks. A companion analysis by our group, currently under review, indicates that in large measles outbreaks, there may be a substantial number of unreported cases (4).

Over the past decade, vaccination rates in the United States have declined significantly, resulting in a sharp increase in measles cases (2,5). In January 2025, a large measles outbreak began in Texas (6), and subsequently, many other US states began seeing an increase in measles cases. In 2025, a total of 2,287 confirmed measles cases were reported across 44 states, and over 2,100 confirmed cases were reported in the first six months of 2026 (7). As measles cases surge, understanding the transmission dynamics of measles is key, not only to responding to and preventing localized outbreaks, but also controlling spread around the globe.

In Utah, measles cases first emerged in June 2025, and a large regional outbreak began in August 2025 in Northwest Arizona/Southwest Utah (8). While cases in the state initially remained largely restricted to Southwest Utah, other areas of the state saw large increases in cases in 2026 (9). Traditional epidemiological methods in response to measles outbreaks include contact tracing of social networks like households and schools, but hesitancy within the communities to share information with public health officials limited the ability to successfully map transmission networks throughout the state.

In June 2025, the Utah Public Health Laboratory (UPHL) began performing whole genome sequencing (WGS) on clinical measles virus samples from both Utah and Arizona. Traditional epidemiologic investigations utilize genotyping based on the N450 region, and WGS has generally been used to assess measles outbreaks only retrospectively (10–12). However, WGS allows for a greater resolution when assessing transmission dynamics. UPHL was able to perform WGS in near real-time as the outbreak progressed, and this data, alongside traditional epidemiologic data, provided a powerful tool for resolving transmission networks and responding to measles outbreaks. As cases spread throughout the state, WGS data became critical to overcome the limitations of traditional contact tracing when investigating measles transmission.

## Materials and Methods

### Laboratory Sequencing

UPHL performed whole genome sequencing (WGS) on PCR-positive measles specimens, which comprised nasal or throat swabs, serum, and other unknown sources. To maximize sample acquisition, UPHL leveraged established partnerships with local clinical diagnostic facilities and national reference laboratories (e.g., LabCorp, Quest) for specimen referral. Consequently, UPHL sequenced all internally-tested positive specimens and the majority of externally-tested specimens.

Samples were batched weekly, averaging four days from receipt to sequencing. Automated liquid handling and tiling amplicon library preparation allowed multiplexing of measles, SARS-CoV-2, influenza, and RSV specimens on a single Illumina NextSeq 2000 run. Sequencing was performed at the end of each week, which allowed for immediate bioinformatic analysis and epidemiological integration by the following Monday.

### Bioinformatic Analysis

Total nucleic acid was extracted from measles positive specimens using the Applied Biosystems MagMAX Viral/Pathogen Nucleic Acid Isolation Kit on the KingFisher Flex platform. Sequencing libraries were prepared from this nucleic acid using Illumina’s COVIDSeq RUO Library prep kit, substituting the SARS-CoV-2 primer set with the Artic Measles 400 bp v1.0.0 primer set. The resulting libraries were then sequenced on an Illumina NextSeq 2000 sequencer (13,14). The resulting sequencing data was processed using the Cecret bioinformatic workflow (https://github.com/UPHL-BioNGS/Cecret), which includes the following analysis steps.

Only reads between 400 bp and 700 bp were included. Reads were cleaned using SeqyClean with the default quality cut-off of 20. Cleaned reads were aligned using BWA to the RefSeq reference genome (NC_001498). Primers were trimmed using iVar. Finally, iVar was used to generate consensus sequences where bases must have a minimum quality of 20, a minimum depth of 100, and a minimum frequency of 60% in order to be called. Sequences were genotyped using Nextclade. Only genotype D8 samples with at least 90% coverage in consensus sequences and less than 75% ambiguous bases were included in the analysis.

### Phylogenetic Analysis

We used BEAST2 to create time-scaled phylogenetic trees, which requires a multiple sequence alignment and dates associated with each sequence as inputs (15). Case dates for each sequence were either the disease onset date recorded in the UT-NEDSS (EpiTrax) system, Utah’s disease surveillance system, or, in cases where this date was unavailable, the sample collection date was used as a proxy. ClustalW alignments were created using Clustal Omega, where they were aligned to the D8 reference genome (NC_001498) (16). These alignments were imported into Beauti (15) along with the case dates. We assume an HKY substitution model (17), a strict molecular clock with a rate of 6.61e^-4^ (18), and a coalescent constant size posterior. A maximum clade credibility tree was generated using the TreeAnnotator application (19). Maximum likelihood trees were also generated from the ClustalW alignments using iqtree3 with an HKY substitution model (17,20).

We used these phylogenetic trees to identify monophyletic clades corresponding to distinct introductions of measles into Utah. A sustained introduction was defined as an introduction where there were more than five cases associated with the clade. A limited introduction was defined as an introduction consisting of five cases or fewer that were associated with the given introduction clade.

### Genomic Cluster Analysis

SNP distance matrices were generated using the snp-dists package (21) in Python. For the purposes of these matrices, ambiguous bases were not counted. We used the transcluster package in R to define genomic clusters (22). This model uses SNP distance, temporal distance, and R_0_ to estimate the number of intermediate transmissions there may have been between two cases, and clusters cases together based on a user-defined threshold of intermediate transmissions. We calculated the lambda and beta parameters for this model with the following formulas, using a substitution rate 0f 6.61e^-4^ and a serial interval of 11.9 days (18,23).

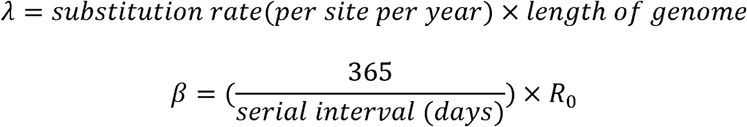

To determine the proper transmission threshold and R_0_ for these genomically-defined clusters, we compared them against contact-based clusters, defined as groups of individuals who had a confirmed, direct exposure to one another, as identified through standard public health case interviews. This comparison is to ensure our genomic-based clusters are capturing the closest true approximation of direct transmissions from the contact-based clusters, but are still able to include other cases based on genomic relatedness. To do this, we calculated the Adjusted Rand Index between contact-based clusters and transmission clusters created from transmission thresholds between 0-9 intermediate transmissions and R_0_ between 1-18. Using this metric, we determined that it was most appropriate to create transmission clusters with a transmission threshold of eight and an R_0_ of five.

Using these transmission clusters, we assessed how well these transmission clusters aligned with epidemiological information. When possible, state and local epidemiologists performing case investigations collected data on school affiliation (defined as attending or working at a specific educational institution). To assess the concordance between transmission clusters and school affiliation, we fit a generalized linear mixed model, accounting for spatial dependence across case pairings. Separately, we also evaluated the proportion of individuals who were unvaccinated in each identified transmission cluster, based on the vaccination status recorded by case investigators.

### Epidemiologic Cluster Analysis

Epidemiologic metadata was collected for Utah samples when possible by state and local epidemiologists and accessed from the UT-NEDSS (EpiTrax) system. Metadata included demographic and geographic variables, vaccination status, and data about potential exposure locations. We investigated possible transmission clusters based on either genomic similarity or shared epidemiological exposures, including workplace, educational, or extracurricular affiliations, as these settings represent environments with prolonged indoor congregation known to facilitate airborne measles transmission (24). We annotated phylogenetic trees with epidemiologic metadata to assess the concordance between molecular data and traditional surveillance. Specifically, we focused on genomic clusters with high diversity in geographic location or institutional affiliation to reconstruct potential transmission patterns throughout the state and within specific social networks. All statistical analyses and visualizations were performed in R version 4.5.1.

### Ethical Approval

This study was conducted in accordance with the Declaration of Helsinki. As this study used publicly available pathogen genomic data and anonymized patient demographic data, formal ethical approval was not necessary.

## Results

Between June 13, 2025 and June 13, 2026, there were 687 confirmed measles cases in Utah. The Utah local health districts with the highest per capita rate of measles cases were Southwest Utah, TriCounty, and Central Utah (Figure 1).

**Figure 1:**
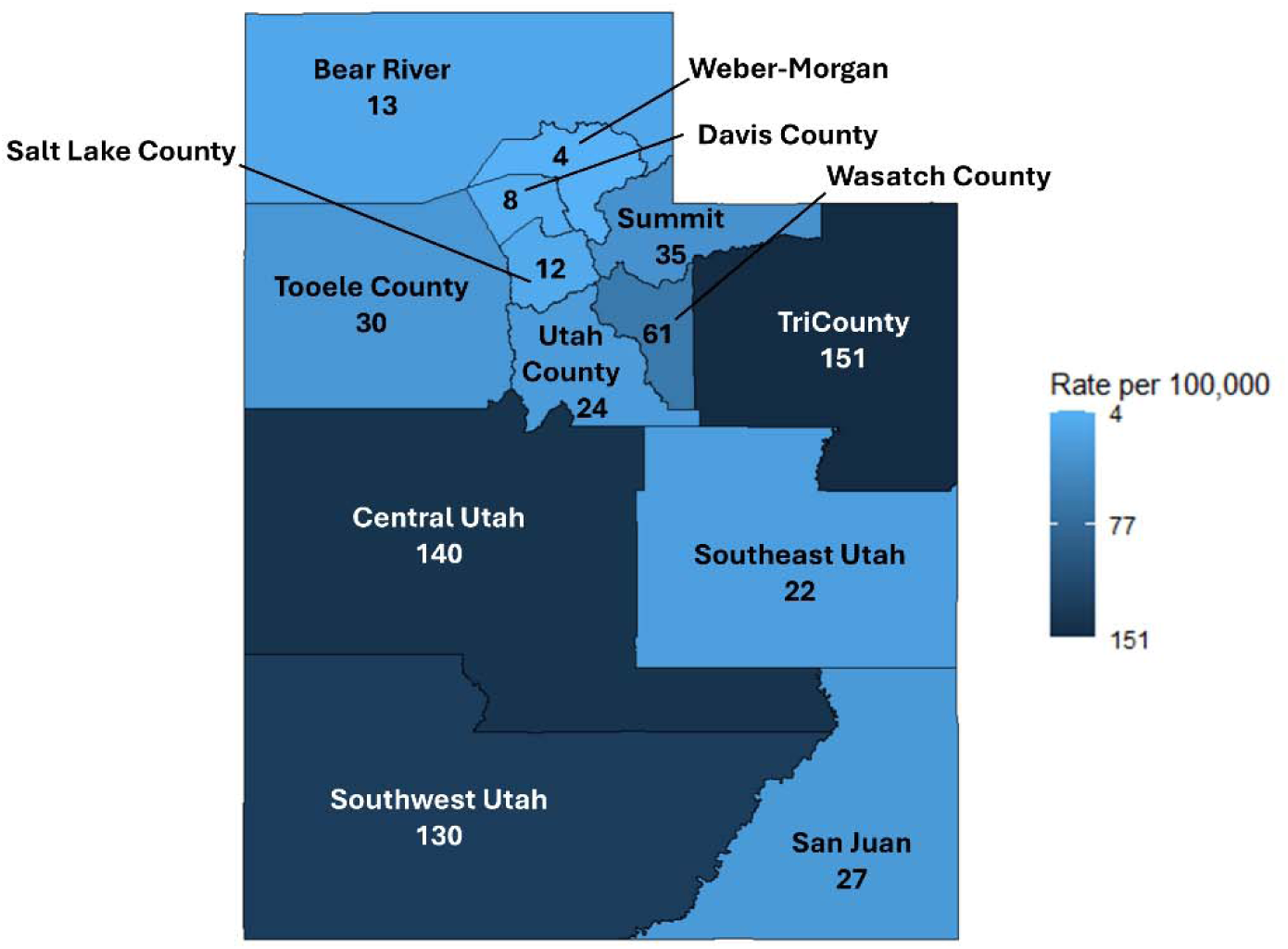
Rate of measles cases per 100,000 residents from June 13, 2025 to June 13, 2026, labelled and colored by rate for each local health jurisdiction in Utah. Rates are calculated using local health jurisdiction population levels reported from the Utah Department of Health and Human Services, Indicator-Based Information System for Public Health website: https://ibis.utah.gov/ibisph-view/.

The median age of all measles cases in Utah was 14. Among cases with recorded race and ethnicity data, 76.4% identified as White, non-Hispanic (n = 525), compared to about 72% in the overall Utah population. However, missing demographic data in 16.3% of cases limits direct population comparisons. The most common jurisdiction of residence for all measles cases was Southwest Utah (n = 272, 39.9%), followed by Utah County (n = 113, 16.6%). However, sequencing coverage varied widely across the state as a result of different rates of testing and healthcare seeking behavior throughout the state. Compared to measles cases that were not sequenced, sequenced cases were more likely to reside in Southwest Utah (46.5% of sequenced cases). These characteristics are summarized in Table 1.

**Table 1:** Demographic characteristics of all measles cases in Utah (June 13, 2025 - June 13, 2026) and all sequenced measles cases in Utah during the same time period.

|  | **Sequenced UT Measles Cases**<br>**N (%)** | **Not Sequenced UT Measles Cases**<br>**N (%)** |
|---|---|---|
| **Sex** |  |  |
| Male | 217 (47.8%) | 117 (50.2%) |
| Female | 229 (50.4%) | 95 (40.8%) |
| Unknown/ missing | 6 (1.3%) | 21 (9.0%) |
| **Race and ethnicity** |  |  |
| White alone, non-Hispanic | 358 (78.9%) | 186 (79.8%) |
| Hispanic or Latino (any race) | 22 (4.8%) | 2 (0.9%) |
| Some other race alone, non-Hispanic | 29 (5.5%) | 7 (12.9%) |
| Unknown race, non-Hispanic | 45 (9.9%) | 15 (6.4%) |
| **Age** |  |  |
| Median (IQR) | 16 (15) | 12 (10) |
| **Has the patient ever received a measles-containing vaccine?** |  |  |
| Yes | 53 (11.7%) | 10 (4.3%) |
| No | 376 (82.8%) | 214 (91.8%) |
| Unknown | 25 (5.5%) | 9 (3.8%) |
| **Jurisdiction** |  |  |
| Central Utah | 24 (5.2%) | 64 (27.5%) |
| Salt Lake County | 42 (9.3%) | 20 (8.6%) |
| Southwest Utah | 211 (46.5%) | 61 (26.2%) |
| TriCounty | 47 (10.4%) | 35 (15.0%) |
| Utah County | 74 (16.3%) | 39 (16.7%) |
| All other jurisdictions | 56 (12.3%) | 14 (6.0%) |

### Laboratory Turnaround and Real-time Utilization of Genomic Data

UPHL performed whole genome sequencing on 454 positive measles specimens (66% of total cases in Utah), including nasal or throat swabs (81%), serum (3%), and unknown sources (16%). On average, the specimen transit time from sample collection to receipt at UPHL took seven days. Multiplexing measles, SARS-CoV-2, influenza, and RSV specimens, combined with quarter-volume reactions, reduced sequencing costs to below $90 per sample.

Overall, this laboratory pipeline yielded an average turnaround time of two weeks from sample collection to the delivery of genomic data for public health interpretation. Incomplete clinical capture persisted because not all symptomatic individuals sought testing and not all tested specimens were sequenced; however, the timeline and volume of WGS for this measles outbreak enabled near-real-time integration of genomic data into epidemiological response efforts. Genomic data was used by epidemiologists to gain insights into the nature of the outbreak, including the number of introductions of measles into the state. The addition of genomic data during case investigations also supplemented the epidemiological information gathered during case interviews, or provided additional insight if epidemiological data was not available due to loss to follow-up. Epidemiologists utilized genomic data in some situations to rule out linkage to known clusters, or suggest potential associations with other cases, thus informing their ongoing surveillance and case investigation.

### Isolated and Sustained Measles Introductions

Using WGS data, we identified a total of five distinct introductions of measles into Utah since June 2025, including two that resulted in sustained transmission and three more that were considered isolated introductions, resulting in five or fewer cases each.

The first sustained introduction occurred in June 2025 in a case from Utah County with known travel history to Southwest Utah. Genomic evidence indicated that this is the first known case linked to the wider Utah-Arizona border outbreak, which sustained transmission through at least June 2026, accounting for 95.6% of cases in the first 12 months of the outbreak. The second sustained introduction occurred in December 2025 in a case from Salt Lake County. Genomic evidence linked this introduction to 20 cases in Utah and six cases in Arizona. While this introduction resulted in sustained transmission, we did not identify new cases linked to this introduction after March 2026.

In addition to these introductions that lead to substantial transmission, we identified three isolated events that did not sustain continued transmission. The first of these introductions occurred in June 2025, including five confirmed cases with close contact. These cases originated in Utah County, and investigators did not identify a clear exposure source. Specimens collected from two of these five cases were sequenced, and subsequent genomic evidence suggested that there was no further transmission from this small outbreak in Utah (Figure 2A). The second isolated introduction occurred in December 2025 among four cases with close contact. These cases originated in Bear River, and investigators were also unable to identify a definitive exposure source. A specimen collected from one of the four cases was sequenced, and genomic evidence suggested that this introduction was not related to any of the ongoing outbreaks at the time, and no further transmission occurred from these cases (Figure 2A). The third isolated introduction involved a single Salt Lake County case in January 2026 with no known exposure sources. Genomic evidence showed this infection was unrelated to ongoing outbreaks and resulted in no further transmission (Figure 2A).

**Figure 2:**
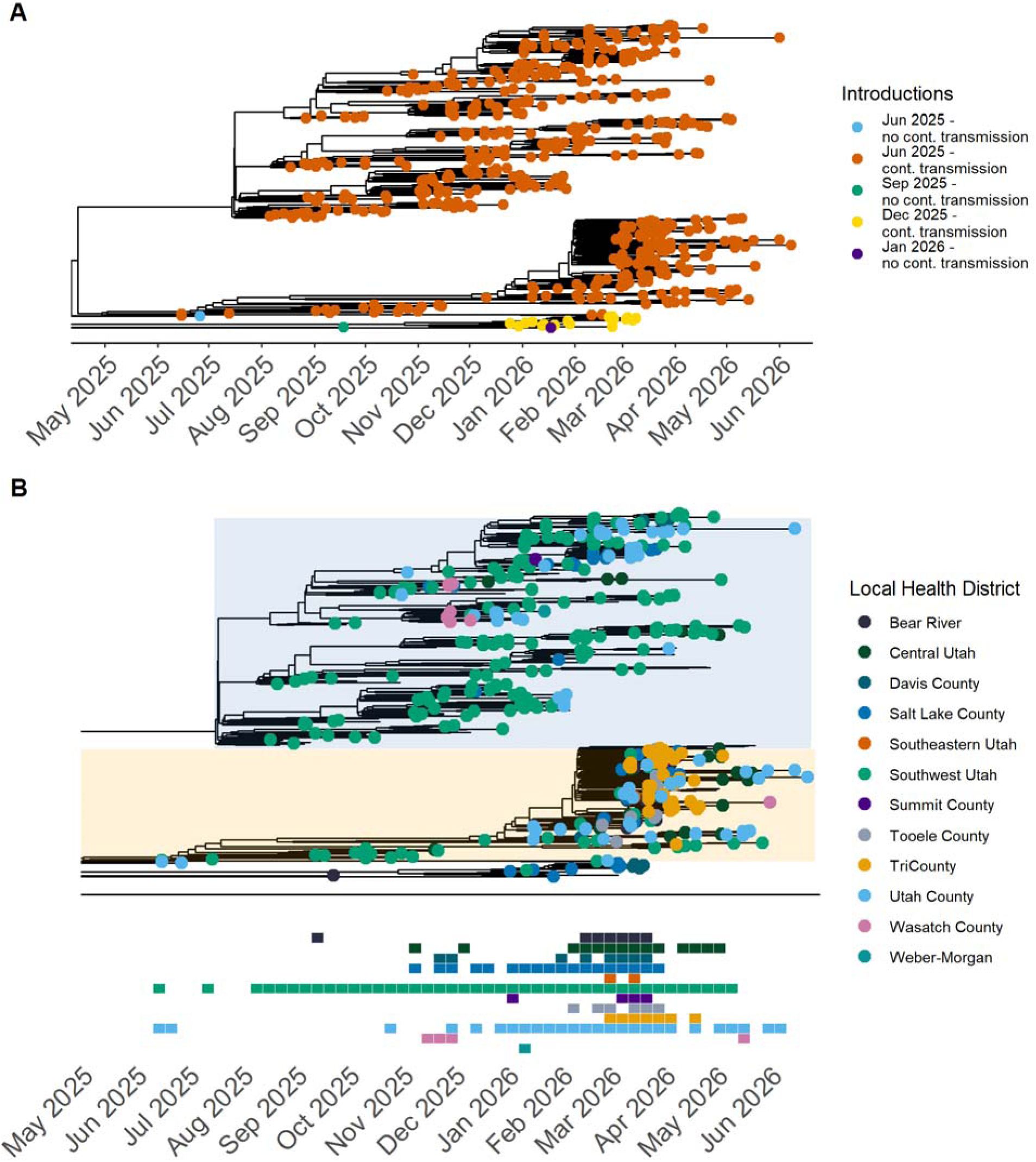
(A) A time-scaled phylogenetic tree with the tips representing all cases recorded in Utah. Tips are colored by cases associated with different introductions of measles into Utah. Whether transmission from a given introduction is isolated or not is labelled in the legend. (B) A time-scaled phylogenetic tree with tips representing each case recorded in Utah. Tips are colored by local health jurisdiction. Below, a heatmap of when cases appeared in each local health jurisdiction on the same temporal axis as the phylogenetic tree.

When there is no identifiable exposure derived from a contact tracing investigation, it is difficult to discern whether a case is part of an ongoing outbreak, especially if the patient does not live or have any travel history to the location of the outbreak. In these instances, genomic data allowed us to determine which cases were likely new introductions, and which were likely part of the larger outbreak. Moreover, genomic data showed that only two of these clades resulted in statewide transmission, while the others were contained to five or fewer cases, providing epidemiologists with improved situational awareness of the outbreak.

### Statewide Spread of Distinct Lineages

Of the five distinct introductions of measles, the first sustained introduction from June 2025 resulted in the most substantial transmission, which ultimately spread statewide. However, genomic evidence suggests that the geographic spread of measles was different in 2025 compared to 2026.

While the first cases in Utah initially emerged in Utah County, the majority of early cases in this outbreak were concentrated in Southwest Utah. Transmission mostly occurred in Southwest Utah through the end of 2025, with 73% of cases in that year occurring there and 27% of cases occurring in other local health districts (Figure 2B). However, transmission patterns shifted in 2026, with cases emerging in all 13 local health districts in Utah. In 2026, only 29% of cases occurred in Southwest Utah, while 71% occurred in other local health districts (Figure 2B).

Notably, the onset of state-wide transmission in February of 2026 coincided with the emergence of two distinct phylogenetic clades. While cases continued to appear in both clades, they were concentrated in different areas of the state. The first clade (shaded in blue), initially identified in Southwest Utah, accounted for 70% of the cases in that jurisdiction, along with 15% of cases from Utah County, 7% of cases from Salt Lake County, and 8% across other jurisdictions from June 2025 to June 2026 (Figure 2B). In contrast, the second clade (shaded in orange) was first identified in Utah County. This clade comprised a much smaller proportion of cases from Southwest Utah (19%), consisting primarily of infections from TriCounty (27%), Utah County (19%), and Central Utah (12%) (Figure 2B). It is important to note that some jurisdictions, such as Central Utah and TriCounty, had high per-capita rates of measles cases but a low proportion of sequenced cases, potentially resulting in under-representation of these jurisdictions in phylogenetic analyses. However, in general, the formation of these two distinct clades highlighted the differences in transmission patterns between the first and second halves of the outbreak.

### Transmission Within and Between Schools

As measles spread throughout the state, schools and childcare settings were notable to epidemiologists as potential sources of transmission. In order to evaluate the potential role of schools and childcare settings in disease transmission, we used genomically-defined transmission clusters to assess whether cases with a shared school affiliation also shared plausible transmission links. We identified 208 potential transmission clusters using genomic data, of which 190 (91%) were composed entirely of unvaccinated individuals. We found a significant association between transmission cluster and school affiliation (odds ratio (OR) = 10.07 95% credible interval (CI) 8.77, 11.55). Figure 3 shows the school affiliation, if any, of each case in the phylogenetic tree. If a school is affiliated with fewer than 10 cases, it is not listed here. Generally, cases from the same school clustered together genomically. We observed notable exceptions in schools located in Southwest Utah, where there was substantial community transmission.

**Figure 3:**
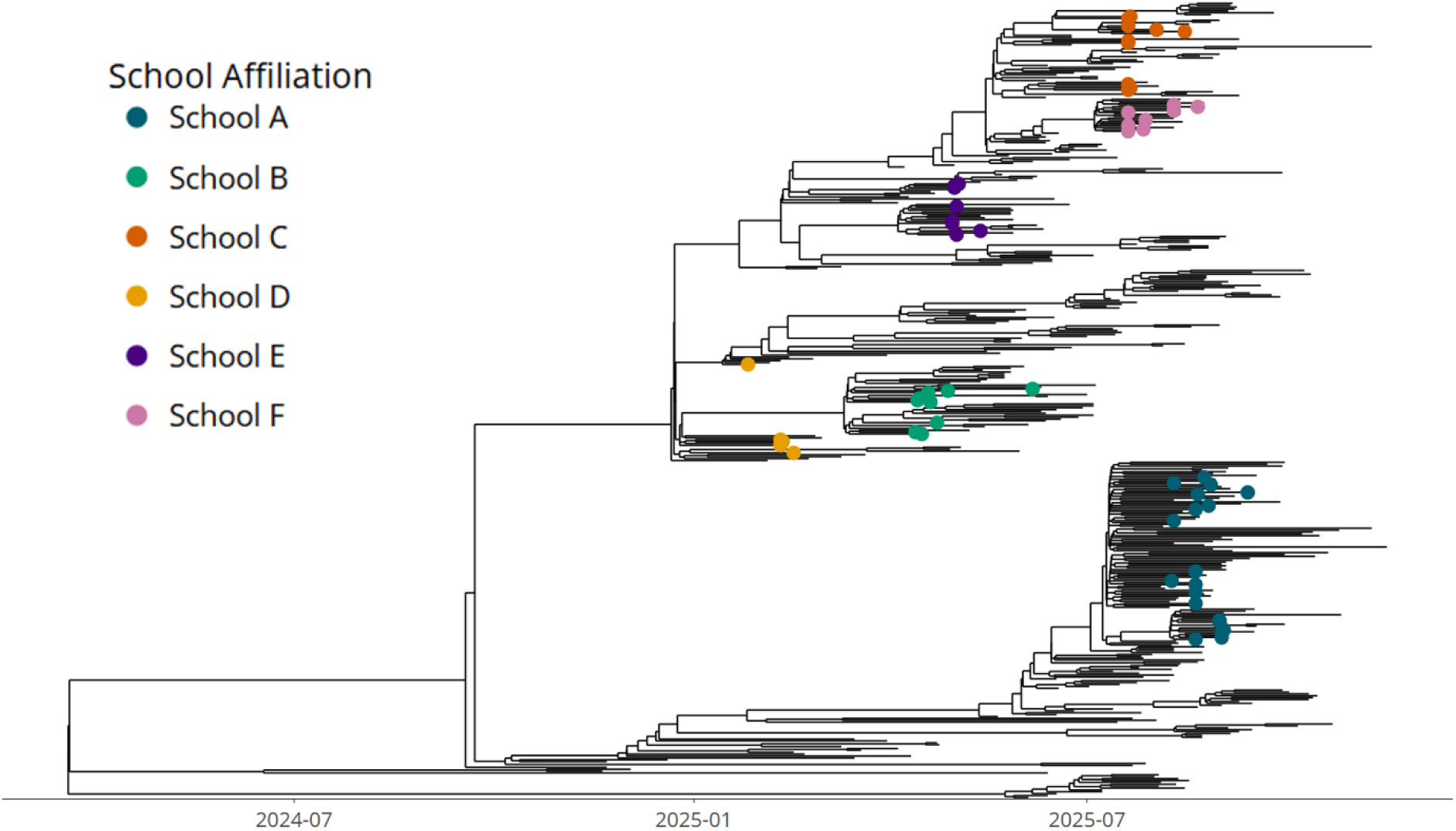
Time-scaled phylogenetic tree including all sequenced Utah measles cases as of June 13, 2026. Tips are colored based on their school affiliation for cases attending a school that was associated with 10 or more measles cases.

In addition to school itself, some youth extracurricular activities stood out as potential drivers of transmission. Beyond localized school-based clusters, we also evaluated the potential role that youth sporting and social events play in measles transmission. We identified notable transmission from a high school wrestling tournament in Orem, Utah in February 2026. As of March 9, six measles cases were identified in boys aged 15–18 who competed in the wrestling event. These cases clustered together in a monophyletic group shared by cases associated with community transmission in Southwest Utah (Figure 4). By March 18, this monophyletic group included 28 wrestling competition attendees, five non-attendees who were known to have close contact with individuals at the competition, and eight others who did not attend the event nor have any known epidemiological connection to student wrestlers. These individuals with no known epidemiological connection included people from across the state, including Southwest, Salt Lake County, TriCounty, and Bear River health jurisdictions. However, it is worth noting that due to the genomic stability of measles, these cases may be genomically similar but epidemiologically distinct, or there may be unsampled intermediate cases, rather than a direct link to the wrestling event.

**Figure 4:**
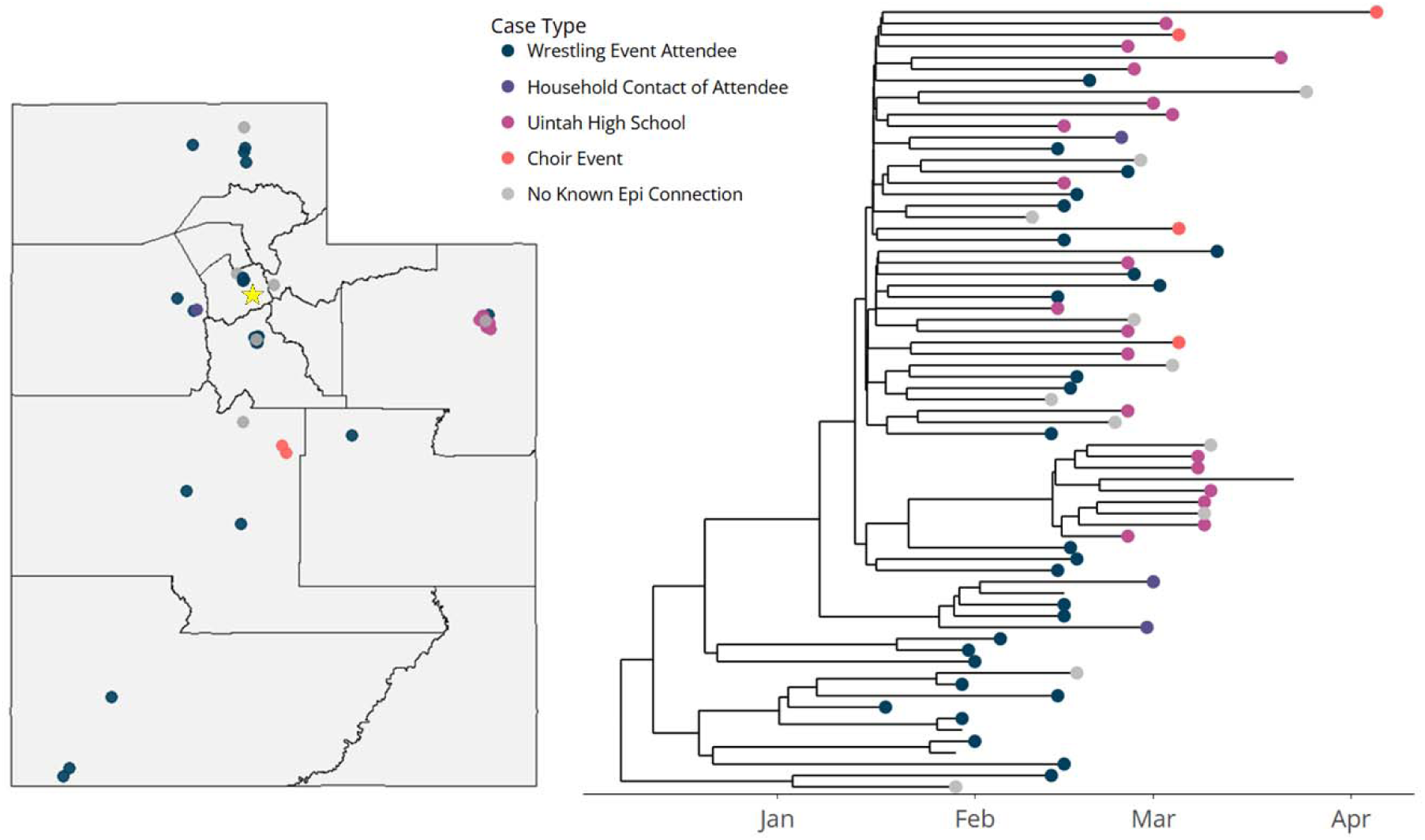
Time-scaled phylogenetic tree of cases associated with a high school wrestling event in Orem, Utah in February 2026. Cases are colored by their epidemiologic association (or lack thereof) with the wrestling event. The location of the wrestling championship event is marked on the map by a star.

As of the end of March, genomic and epidemiologic evidence suggested that cases from this wrestling event may have seeded secondary outbreaks in different parts of the state. Notably, a secondary outbreak that began in TriCounty Health District at the end of February was likely seeded by student athletes attending Uintah High School who competed in the wrestling event. By early April, there were a total of 25 cases affiliated with Uintah High School, all of which clustered genomically with cases from the wrestling event. These likely represent at least a second generation of transmission from the wrestling event, as these cases had no known specific connection to this event or student athletes, other than simply attending Uintah High School.

Similarly, genomic evidence suggests potential continued transmission from this cluster in late March in a choir event in Central Utah. Four individuals involved in this event became ill with measles, and all of these cases were shown to be genomically similar to cases from the wrestling outbreak. However, while the genomic evidence suggests a potential transmission link, there is no documented social or geographic connection, and another explanation, such as an unsampled intermediate case, is also plausible, especially considering the low rate of testing and sequencing in Central Utah.

Overall, the monophyletic group of cases associated with the wrestling event is one of the larger clades in this outbreak, including 69 cases as of April, 2026. Of these, 56 (81%) were unvaccinated or had unknown vaccination status. Cases in this monophyletic group include individuals associated with at least ten different schools throughout the state, located in at least eight of the 13 local health jurisdictions in Utah. While it is not possible to establish a definitive connection between cases based solely on genomic similarity, the genomic evidence provides insight into the transmission that may have followed the wrestling event, which was not possible from traditional epidemiological data alone.

We identified another cluster that occurred in early February 2026. The initial cases in this cluster resided and attended elementary, middle, or high schools in Southwest Utah, including several students who participated in their school’s basketball teams (Figure 5A). Over multiple weeks, this cluster expanded, both within Southwest Utah and throughout the state (Figure 5B). By February 19, it included cases in Salt Lake County and Utah County that all had a known connection to high school basketball games, suggesting that these events may facilitate transmission. However, in early March, cases emerged in this cluster that were not linked to basketball (Figure 5C). Several secondary clusters developed within this cluster, including at least eight cases associated with a preschool in Southwest Utah, six cases associated with a private school in Salt Lake County, and four cases in Utah county who were all associated with music and dance extracurriculars. By March 9, this monophyletic group included 53 cases from Southwest Utah, Salt Lake County, Utah County, and Summit County, 94% of whom were unvaccinated or had unknown vaccination status.

**Figure 5:**
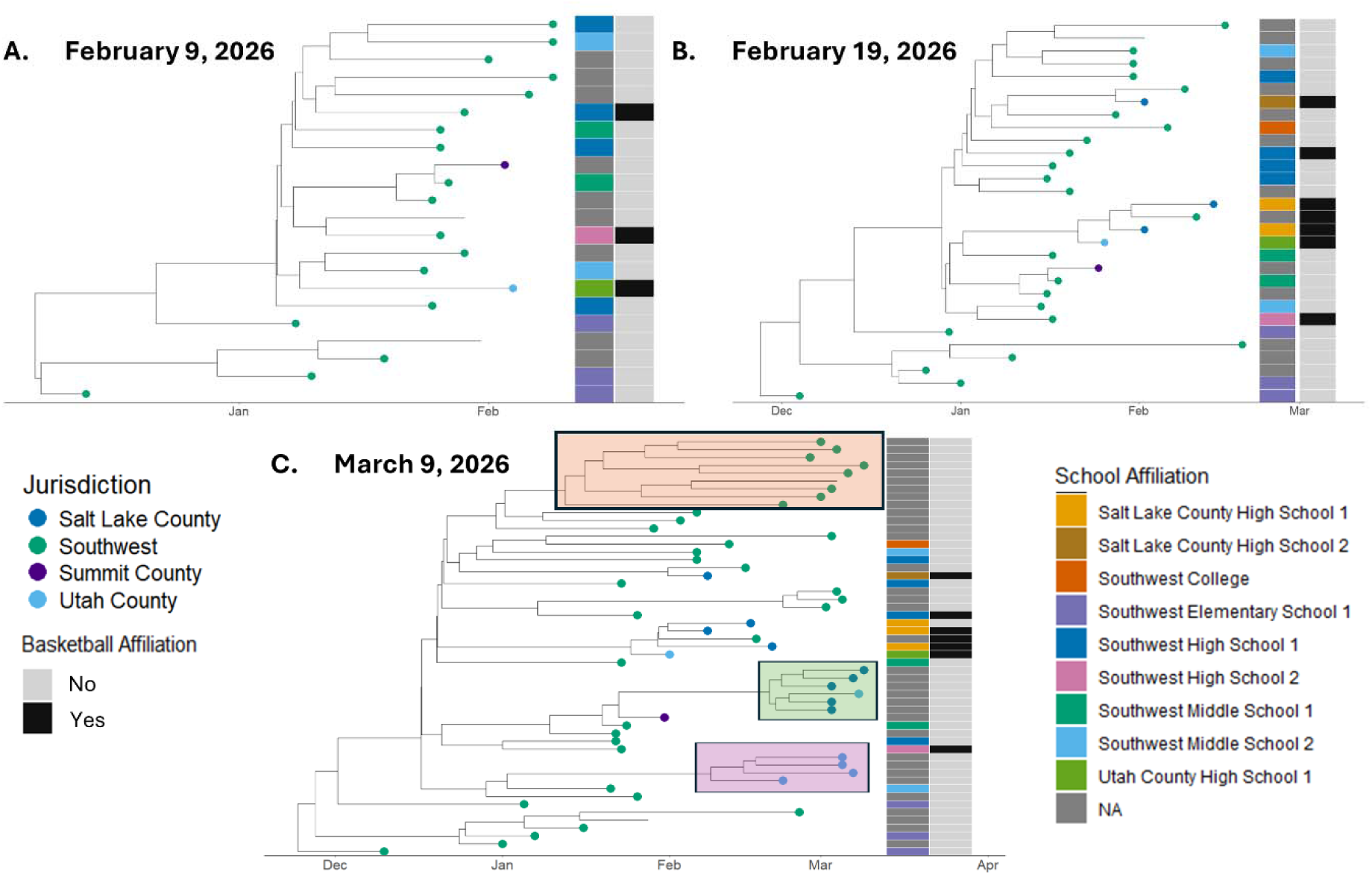
(A) Time-scaled phylogenetic tree showing a genomically-related cluster of cases as of February 9, 2026. Tips are colored based on the jurisdiction of residence of the patient, and colored bars represent the school affiliation (if any) of the patient, and whether the patient was associated with a high school basketball team. (B) Time-scaled phylogenetic tree showing the same genomically-related cluster of cases as of February 19, 2026. Tips are colored based on the jurisdiction of residence of the patient, and colored bars represent the school affiliation (if any) of the patient, and whether the patient was associated with a high school basketball team. (C) Time-scaled phylogenetic tree showing the same genomically-related cluster of cases as of March 9, 2026. Tips are colored based on the jurisdiction of residence of the patient, and colored boxes highlight subclusters of interest that are associated with music/dance extracurriculars (purple), a private school in Northern Utah (green), and a daycare in Southwest Utah (orange).

Lastly, we also evaluated a transmission cluster associated with a statewide youth mountain biking race in Wasatch County in August 2025. Despite widespread exposure to an infectious participant, only five subsequent measles cases were epidemiologically linked to the event, all residing in Southwest Utah. Genomic sequencing of three available specimens revealed that only two (Case A and Case B) shared a monophyletic group, suggesting a likely transmission chain (Figure 6). The third sequenced case (Case C) was genomically distant, suggesting it may not be linked in a transmission chain with the bike race cases.

**Figure 6:**
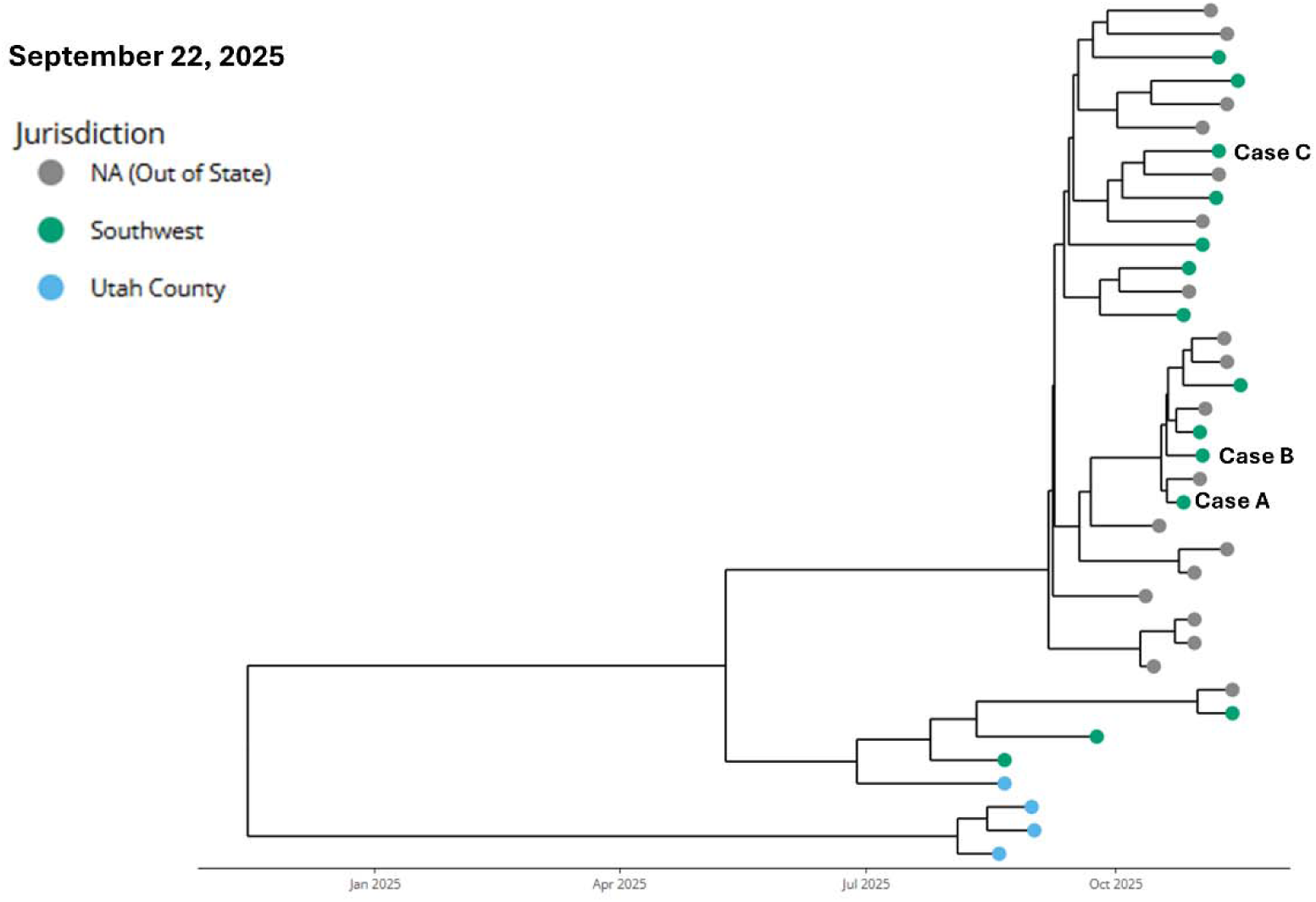
Time-scaled phylogenetic tree including all sequenced Utah measles cases between January 1, 2025 through September 22, 2025. Three sequenced cases associated with the youth mountain biking event are labelled Case A, B, and C. Tips are colored based on their jurisdiction of residence within Utah.

## Discussion

This study demonstrates the utility of genomic analyses in the context of disease outbreaks and highlights their ability to enhance epidemiologic data. The integration of genomic and epidemiologic data provided more insight into the transmission dynamics of the outbreak, helping public health professionals better understand the transmission patterns throughout the state. We were able to generate genomic-based transmission clusters, informed by epidemiological data, to help fill in the gaps left by traditional surveillance methods alone. These transmission clusters highlighted that cases with shared school affiliation were more likely to be genomically related. Examining specific phylogenetic clusters emphasized that schools and youth activities played a key role in the transmission of measles throughout the state, with certain high risk indoor sports, such as basketball and wrestling, being associated with these large disease clusters.

Our findings are consistent with other studies that highlight the utility of WGS data in understanding measles outbreaks and illuminating specific transmission factors (18,25–28). For example, Masters et al. used genomic data to resolve transmission chains involved in a measles outbreak among Afghan refugees involved in Operation Allies Welcome in 2021 (25). WGS data showed that multiple genomically distinct measles lineages were imported in this event, and some epidemiological groupings contained cases representing multiple viral lineages. Thus, they found that epidemiologic cluster identification could be further refined by the addition of genomic analyses. Hiebert et al., found WGS data to be similarly helpful in analyzing the measles outbreaks occurring in Canada over a three year period, and were able to use genomic data to rule out undetected links between cases and place cases in distinct clades (18).

Specifically, WGS offers improved resolution over genotyping or N450 sequencing (29). The global genomic diversity of measles has declined in recent years and only two genotypes are known to be circulating (29). Currently, 90% or more of cases in North America belong to the D8 genotype, while a smaller minority belong to the B3 genotype (2,30). Because of this, traditional genotyping does not offer a high degree of clarity for resolving transmission chains (29). In contrast, WGS facilitates better discrimination of measles variants and surveillance of transmission dynamics (26). This granularity is particularly important when evaluating whether an outbreak is driven by multiple distinct introductions or by a single prolonged chain of local transmission. This level of detail was useful in Israel, which experienced a measles outbreak during 2018–2019. WGS data revealed that this outbreak actually consisted of several distinct clusters sparked by separate introduction events that occurred at different time points during the outbreak (11). Conversely, in the case of Utah’s outbreak, while there have been multiple distinct introductions, our WGS analysis shows that the most widespread of these introductions has been circulating continuously in the state since the initial outbreak was identified.

The utilization of genomic data can also enhance surveillance and response efforts by influencing public health messaging and potentially informing more effective interventions. For example, our investigation suggests that youth sports and extracurriculars can play a key role in fueling transmission and introducing the virus to new populations, but the risks associated with these activities vary significantly based on the nature and setting of the event. Specifically, the youth mountain bike race was a large event with many participants from throughout the state, but despite the size of the gathering, there were relatively few cases that appeared to be linked to that event, based on genomic evidence.

In contrast, the cluster associated with high school basketball seems to demonstrate how a sporting event like basketball, which is played indoors and with more potential close contact than biking, could accelerate an outbreak that is circulating in a particular region. Initially, this cluster appeared to be driven by community transmission in Southwest Utah, with many genomically related cases in children from different schools, but when high school basketball teams traveled throughout the state to play, genomically similar cases appeared in new regions. This genomic pattern suggested that high school basketball may have acted as a means to introduce measles from where it was spreading in Southwest into new communities in the state, potentially causing secondary outbreaks in communities with no known connection to basketball.

The high school wrestling tournament provides an even more salient example of this phenomenon. Students and their families from many areas of the state convened for this competition, which, like basketball, took place indoors and had an even greater level of close contact. Genomic and epidemiological data suggest that many participants were infected at the competition and brought measles home with them, introducing it into new communities. This is one of the larger and more complete genomic clusters in our surveillance, and it offers insight into how a sporting event can accelerate disease transmission throughout the state and potentially trigger secondary outbreaks, even in the absence of confirmed epidemiologic connections.

The analysis of these clusters can help public health staff target interventions to prevent the spread of disease in future outbreaks. For example, 91% of genomically-identified disease clusters were comprised entirely of individuals who were unvaccinated, and the clusters associated with youth sports, like wrestling and basketball, also had low rates of vaccination.

WGS data can be used to strategically choose the most critical issues to address in order to maximize prevention. For example, genomic surveillance may be a useful tool for informing stakeholders about the risks of different types of events, or identifying specific populations, such as youth participating in sports, who may benefit from targeted vaccination efforts. This is particularly important in situations with limited resources, or where it may be difficult to implement public health interventions at the local level.

In addition to providing insight into transmission patterns, the cluster of cases associated with high school basketball also provides evidence for missed cases and undetected transmission. For example, one young adult may have been missed using epidemiologic data alone, as they did not attend school nor any basketball games, but genomic data showed their infection was closely linked to the virus circulating amongst several basketball players and game attendees. Epidemiologic investigation revealed that this case had a close contact who attended basketball games in northern Utah and returned home sick, but was never tested. In this situation, the integration of genomic and epidemiological data for surveillance improved our understanding of the potential transmission chain in this situation.

This study is one of the largest genomic epidemiology studies of measles to date, and it shows the utility of integrating genomic and epidemiologic data in near-real-time for enhanced surveillance. Insights from this analysis have implications for approaches to control measles transmission and how to respond to future outbreaks. However, this study also has several limitations. Despite the large sample size and relatively large proportion of sequences available from this outbreak, the data used in this study was an incomplete picture of this outbreak. Not everyone who became ill sought healthcare, not everyone who did was tested, and not every tested sample was sequenced. In particular, some jurisdictions with high case rates but low sequencing rates, such as Central Utah, may be disproportionately excluded from the phylogenomic analysis. The topography of the phylogenetic tree, with long terminal branch lengths, also suggests missing cases. Low genome coverage can also result in missing or poorly resolved loci, impacting phylogenetic interpretations. The addition of this missing data might lead to different interpretations of the dynamics of this outbreak.

Additionally, measles virus shows remarkable genomic stability (18,31), which poses a challenge for genomic epidemiology. With a high degree of genomic stability, there may be cases in a large outbreak that are genomically similar but epidemiologically distinct. However, our study builds upon existing research showing that despite this limitation, genomic data can be used in conjunction with traditional epidemiologic data for improved resolution of transmission dynamics (10,18).

## Conclusions

In conclusion, our findings illustrate the advantages of using WGS data in real-time measles surveillance and response. Genomic sequencing has proven to be a powerful tool to maintain and advance the goal of eliminating measles, especially as the resurgence of measles globally threatens these efforts. Genomic surveillance plays an important role in maintaining a robust surveillance capacity and can lead to better identification of and response to measles outbreaks.

## Data Availability

All genomic data are publicly available under BioProject PRJNA1293457

https://www.ncbi.nlm.nih.gov/bioproject/PRJNA1293457

## Declarations

### Availability of data and materials

All genomic data are publicly available under BioProject PRJNA1293457.

### Authors’ contributions

MJ performed epidemiologic cluster analysis, performed data visualization, and co-wrote the original draft of the manuscript.

AM performed bioinformatic and phylogenetic analysis, performed data visualization, and co-wrote the original draft of the manuscript.

CN contributed to data management and provided critical revisions or the manuscript.

LN provided critical revisions of the manuscript and insights into analysis.

AS contributed to data management and provided review and editing of the manuscript. KO contributed to supervision and provided review and editing of the manuscript.

All authors read and approved the final manuscript.

## Acknowledgements

We would like to thank the Utah Public Health Lab Sequencing and Bioinformatics team, including Nick Madej, Katie Maguire, Jenni Wagner, and Robby Sainsbury. We would also like to thank the epidemiologists throughout the state of Utah who worked to gather epidemiological data about measles cases for surveillance and contact tracing.

## References

1. History of Measles [Internet]. CDC; 2026. Available from: https://www.cdc.gov/measles/about/history.html#:~:text=400%20to%20500%20people%20died,encephalitis%20(swelling%20of%20the%20brain)

2. Mathis AD, Clemmons NS, Redd SB, Pham H, Leung J, Wharton AK, et al. Maintenance of Measles Elimination Status in the United States for 20 Years Despite Increasing Challenges. Clin Infect Dis. 2022 Aug 31;75(3):416–24. doi:10.1093/cid/ciab979

3. Van Boven M, Kretzschmar M, Wallinga J, O’Neill PD, Wichmann O, Hahné S. Estimation of measles vaccine efficacy and critical vaccination coverage in a highly vaccinated population. J R Soc Interface. 2010 Nov 6;7(52):1537–44. doi:10.1098/rsif.2010.0086

4. Marye A, Toth DJA, Yon GV, Wagoner J, Lofgren ET, Samore MH, et al. Hidden Burden of a Measles Outbreak Revealed by Genomic and Transmission Models [Internet]. 2026 [cited 2026 Jul 29]. Available from: http://medrxiv.org/lookup/doi/10.64898/2026.07.09.26357695 doi:10.64898/2026.07.09.26357695

5. Vaccination Coverage and Exemptions among Kindergartners [Internet]. CDC; 2025. Available from: https://www.cdc.gov/schoolvaxview/data/index.html

6. Confirmed Case of Measles - January 2025 [Internet]. Texas Department of State Health Services; 2025. Available from: https://www.dshs.texas.gov/news-alerts/confirmed-case-measles-january-2025

7. Measles Cases and Outbreaks [Internet]. CDC; 2026. Available from: https://www.cdc.gov/measles/data-research/index.html

8. Measles transmission in Southwest Utah and considerations for early extra MMR dose [Internet]. Utah Department of Health and Human Services; 2025. Available from: https://dhhs.utah.gov/wp-content/uploads/8-26-25-DHHS-measles-HAN.pdf

9. Utah measles outbreak response [Internet]. Utah Department of Health and Human Services; 2026. Available from: https://epi.utah.gov/measles-response/

10. Bodewes R, Maissan C, Van De Nes-Reijnen L, Boter M, Van Den Boom SC, Zwagemaker F, et al. Whole genome sequencing of measles viruses: the benefit for outbreak investigation, The Netherlands, 2013–2014. Virus Genes. 2026 Jun 19. doi:10.1007/s11262-026-02253-8

11. Indenbaum V, Bucris E, Friedman K, Kushnir T, Eliyahu H, Azar R, et al. Measles Sequencing: Lessons Learned from a Large-Scale Outbreak. Viruses. 2025 Jun 27;17(7):913. doi:10.3390/v17070913

12. Genetic Analysis of Measles Viruses [Internet]. CDC; 2024. Available from: https://www.cdc.gov/measles/php/laboratories/genetic-analysis.html

13. A pan Measles morbillivirus scheme. [Internet]. Primalscheme Labs. Available from: https://labs.primalscheme.com/detail/artic-measles/400/v1.0.0/

14. Kent C, Smith AD, Tyson J, Stepniak D, Kinganda-Lusamaki E, Lee T, et al. PrimalScheme: open-source community resources for low-cost viral genome sequencing [Internet]. 2024 [cited 2026 Jul 29]. Available from: http://biorxiv.org/lookup/doi/10.1101/2024.12.20.629611 doi:10.1101/2024.12.20.629611

15. Bouckaert R., Vaughan T.G., Barido-Sottani J., Duchêne S., Fourment M., Gavryushkina A., et al. (2019) BEAST 2.5: An advanced software platform for Bayesian evolutionary analysis. PLoS computational biology, 15(4), e1006650.

16. Sievers F, Wilm A, Dineen D, Gibson TJ, Karplus K, Li W, et al. Fast, scalable generation of high quality protein multiple sequence alignments using Clustal Omega. Mol Syst Biol. 2011 Oct 11;7(1):MSB201175. doi:10.1038/msb.2011.75

17. Hasegawa M, Kishino H, Yano T aki. Dating of the human-ape splitting by a molecular clock of mitochondrial DNA. J Mol Evol. 1985 Oct;22(2):160–74. doi:10.1007/BF02101694

18. Hiebert J, Zubach V, Schulz H, Severini A. Genomic tools for post-elimination measles molecular epidemiology using Canadian surveillance data from 2018–2020. Front Microbiol. 2024 Nov 19;15:1475144. doi:10.3389/fmicb.2024.1475144

19. Drummond, A. J. and A. Rambaut. 2007. BEAST: Bayesian evolutionary analysis by sampling trees. BMC Evolutionary Biology 7:214.

20. Wong TKF, Ly-Trong N, Ren H, Demotte P, Baños H, Roger AJ, et al. IQ-TREE 3: phylogenomic inference software using complex evolutionary models. Tamura K, editor. Mol Biol Evol. 2026 May 1;43(5):msag117. doi:10.1093/molbev/msag117

21. Seemann T, Klotzl F, Page A. snp-dists [Internet]. 2019. Available from: https://github.com/tseemann/snp-dists

22. Stimson J, Gardy J, Mathema B, Crudu V, Cohen T, Colijn C. Beyond the SNP Threshold: Identifying Outbreak Clusters Using Inferred Transmissions. Leitner T, editor. Mol Biol Evol. 2019 Mar 1;36(3):587–603. doi:10.1093/molbev/msy242

23. Klinkenberg D, Nishiura H. The correlation between infectivity and incubation period of measles, estimated from households with two cases. J Theor Biol. 2011 Sep;284(1):52–60. doi:10.1016/j.jtbi.2011.06.015

24. Azimi P, Keshavarz Z, Cedeno Laurent JG, Allen JG. Estimating the nationwide transmission risk of measles in US schools and impacts of vaccination and supplemental infection control strategies. BMC Infect Dis. 2020 Dec;20(1):497. doi:10.1186/s12879-020-05200-6

25. Masters NB, Beck AS, Mathis AD, Leung J, Raines K, Paul P, et al. Measles virus transmission patterns and public health responses during Operation Allies Welcome: a descriptive epidemiological study. Lancet Public Health. 2023 Aug;8(8):e618–28. doi:10.1016/S2468-2667(23)00130-5

26. Probert WS, Glenn-Finer R, Espinosa A, Yen C, Stockman L, Harriman K, et al. Molecular Epidemiology of Measles in California, United States—2019. J Infect Dis. 2021 Sep 17;224(6):1015–23. doi:10.1093/infdis/jiab059

27. Hohan R, Surleac M, Miron VD, Tudor A, Tudor AM, Săndulescu O, et al. Ongoing measles outbreak in Romania: Clinical investigation and molecular epidemiology performed on whole genome sequences. Harapan H, editor. PLOS ONE. 2025 Jan 15;20(1):e0317045. doi:10.1371/journal.pone.0317045

28. Stanley S, Patrick RC, Elkan M, Fink J, Oh B, Sun Y, et al. Genomic epidemiology of two travel-associated pediatric measles viruses within the B3 Lineage. J Clin Virol. 2026 Aug;185:105966. doi:10.1016/j.jcv.2026.105966

29. Moss WJ, Griffin DE. What’s going on with measles? Levy DE, editor. J Virol. 2024 Aug 20;98(8):e00758–24. doi:10.1128/jvi.00758-24

30. Epidemiological Alert Measles in the Americas Region [Internet]. Pan American Health Organization; 2026. Available from: https://www.paho.org/sites/default/files/2026/05/2026-may-29-phe-epialert-measles-engfinal.pdf

31. Schrag SJ, Rota PA, Bellini WJ. Spontaneous Mutation Rate of Measles Virus: Direct Estimation Based on Mutations Conferring Monoclonal Antibody Resistance. J Virol. 1999 Jan;73(1):51–4. doi:10.1128/JVI.73.1.51-54.1999

